# Consensus for Integrating the Point-of-Care Genital InFlammation Test (GIFT) into Sexually Transmitted Infection Management Guidelines: Results from a Two-Round Modified Delphi Survey

**DOI:** 10.1101/2024.10.10.24315280

**Authors:** Eneyi E. Kpokiri, Constance Mackworth-Young, Suzanna Francis, Tania Crucitti, Janneke HHM van de Wijgert, Lindi Masson, Jo-Ann Passmore, Emma M. Harding-Esch

## Abstract

**Background:** Sexually transmitted infections (STIs) and bacterial vaginosis (BV) are frequently asymptomatic in women. They cause genital inflammation and can increase transmission and acquisition of HIV. GIFT (Genital InFlammation Test) is a novel, point-of-care (POC) device under development for detecting genital inflammation in women. We aimed to obtain consensus to inform the development of GIFT and its integration into management guidelines.

**Methods:** We employed a Delphi technique through two rounds of online surveys. Respondents included service providers, health programmers, researchers, and policy makers. Round one questions generated ideas, and round two built consensus on the strategies from round one. Survey sections included demographics, implementation of the test, integration into current guidelines and purpose. Round two survey employed a five-point Likert scale from strongly agree to strongly disagree. Consensus was reached if ≥70% of participants selected strongly agree or agree.

**Results:** We received 28 to the first and 68 to the second round. From both rounds, participants were healthcare providers (41%) or researchers (26%), residing in Africa (57%), Europe (21%) and America (10%). Most participants agreed that GIFT should be used as a screening tool to be followed by confirmatory STI testing before treatment: 75% (round 1), 69% (round 2). There was consensus that populations to benefit most from GIFT were young asymptomatic women (16-24 years) in high HIV prevalence settings, and high-risk women like female sex workers and those with multiple partners of any age. Attributes of GIFT ranked as most important included ease-of-use, stability at room temperature, high diagnostic accuracy and barriers were test stock-outs, complexity of use and high cost,

**Conclusion:** While the Delphi process suggests the purpose of GIFT is as a POC screening tool, factors like supply chain, storage and stakeholder engagement are crucial for its integration into management guidelines.

**Key messages:**

- There is a need to identify BV and STIs in asymptomatic women, however, laboratory diagnostics services are severely limited in most low- and middle-income countries
- If the GIFT test has high diagnostic accuracy, it promises to be a valuable point-of-care screening tool for detecting genital inflammation in asymptomatic women and may be useful to inform the management of women with symptoms.
- It is important to obtain feedback from a variety of stakeholders when considering test design and implementation to increase the test’s utility and likely adoption in policy and routine care.

## Introduction

Sexually transmitted infections (STIs) and bacterial vaginosis (BV) are a significant public health burden worldwide, with an estimated 1 million new cases of curable STIs acquired each day and a BV prevalence of 50-60% in women worldwide.^1^ STIs and BV increase the risk of HIV acquisition and reproductive complications, including chronic pelvic pain, infertility, and poor birth outcomes.^2^ In women living with HIV, the presence of STIs and BV may also increase the risk of mother-to-child HIV transmission and transmission to sex partners, due to an increased viral load in the genital tract area.^3^ Importantly, women with asymptomatic STIs or BV also have a higher risk of genital inflammation on the average than women who do not have an infection,^4^ and these asymptomatic conditions are associated with the same adverse sequelae as symptomatic STIs and BV. Continued efforts are therefore needed to address these conditions, whether symptomatic or asymptomatic, despite current guidelines not recommending BV treatment unless symptomatic or during pregnancy, largely because existing therapeutics have limited long-term efficacy.^5^ It is also thought likely that the high HIV incidence rates in the African region reflect, in part, poor management of STIs and BV generally.^3,6^

Access to laboratory diagnostics to screen or test for STIs and BV in most low- and middle-income countries (LMICs) is limited.^7^ Where diagnostic testing is not possible, the World Health Organization (WHO) promotes the use of syndromic management (treatment based on symptoms and occasionally clinical signs) because this approach is easily implemented, relatively inexpensive and patients are treated immediately.^7^ However, clinical signs and symptoms are poor predictors of STIs and BV, and the majority of women who have a laboratory-confirmed discharge-causing STI/BV are asymptomatic.^8^ Given that STIs/BV are often asymptomatic, inflammatory and untreated, and that genital inflammation increases HIV risk, a test that can detect genital inflammation could improve the reproductive health of women in Africa and other regions where syndromic management is otherwise implemented. Such a test in point-of-care (POC) format would provide a low-cost, user-friendly and rapid solution, with the ability to conduct the test and receive results during a single consultation, enabling prompt clinical decision-making, linkage to treatment and preventing complications from late or poor STI/BV management.^7,9^

The Genital InFlammation Test (GIFT) (https://gift.org.za/) is a prototype rapid lateral flow test that has been developed for the detection of three genital inflammation biomarkers (IL-1α, IL-1β and IP-10) that were previously shown to be associated with the presence of STIs and BV, as well as increased HIV acquisition risk, in South African women.^10^ These biomarkers were subsequently validated in five cohorts from different regions in Africa.^6^ The sample type is a lateral vaginal wall swab, and results are produced in approximately 15 minutes. The lateral flow test format was chosen because of its relatively low technology requirements and potentially low cost. The The performance of the device is currently is currently being evaluated in three countries in Africa (South Africa, Madagascar and Zimbabwe).^11^ The main outcome measures will be performance (sensitivity, specificity, positive predictive value [PPV], and negative predictive value [NPV]) of the GIFT device compared to laboratory-based gold standard nucleic acid amplification testing for STIs (*Chlamydia. trachomatis, Neisseria gonorrhoeae, Trichomonas vaginalis* and *Mycoplasma genitalium*) and BV by Nugent score. The device’s sensitivity and specificity to detect at least one STI/BV that would warrant treatment has not yet been established. However, many factors beyond a test’s diagnostic accuracy inform whether it will be implemented within routine practice and into national guidelines.^12,13^ These include attributes of the test itself (such as run-time and cost) and how the test is used (such as its use case and implementation into clinical pathways).

In order to understand how to further optimise the GIFT device, and how it could be integrated into current STI management guidelines, we conducted a two-round modified Delphi survey with a focus on LMICs where rapid STI/BV aetiologic testing is not widely available.

## Methods

### Questionnaire design

A Delphi consensus technique was used in this study.^14^ The Delphi technique is a structured process for consensus development that involves group communication among a panel of respondents selected for their expertise and involvement on a topic, during which convergence of opinion is sought using multiple rounds of survey questionnaires. This approach is well-established, allowing informed decision-making in areas of research where there is little information, through obtaining feedback and building consensus from expert opinions.^14^ The project study team and scientific advisory board (SAB) discussed the focus of the Delphi and developed relevant questions to inform both survey rounds. Round one included a mixture of closed and open-ended qualitative questions (supplementary information 1). The questionnaire was piloted with 10 stakeholders and members of the SAB to refine the questions before study commencement. Findings of the first round informed the questions and statements included in the second round (supplementary information 2). The round one survey was only available in English, whereas the round two survey was translated into French, Spanish and Portuguese.

### Selection of expert Delphi panel

A combined strategy was used to select the Delphi participants. First, a purposive sample of individuals who had previously contributed to international and national STI management guidelines for LMICs was identified from the acknowledgements section of the WHO published guidelines^15^. Second, experts were identified from guideline-writing committees, and included clinicians who are implementors and end-users of the guidelines, and experts with experience in STI management programmes and policy.^15^ A snowball sampling approach was used to identify and recruit additional participants, from STI management programmes, or of policy-making in LMICs. In addition, the study team and SAB identified individuals, who were in turn asked to suggest names and contact details of other relevant stakeholders at the end of the survey.

### Sample size

There is no consensus on the ideal participant size for a Delphi process.^16^ Within our invited participants, we aimed to cover a broad range of expertise across geographic and socio-economic regions. Based on a similar process undertaken for the development of an international sexual and reproductive health survey instrument,^17^ we aimed to invite approximately 100 individuals in order to recruit at least 50 participants, with efforts made to reach a response rate of 75%, which is suggested for each round to achieve or maintain rigour.^18^

### Survey dissemination

The questionnaire was hosted online using JotForm, with an editable Microsoft Word version sent via email for offline completion to increase the response rate. Participants were sent the link to the survey by personalised email and given two weeks for completion. The round one survey was sent out in March 2022 and the round two survey in August 2022. Two rounds of reminder emails were sent to non-responders and a further one week was allowed for participation. For the second round, we created a QR code with the online survey link, and printed invitation fliers which were then distributed in-person at the International Union against Sexually Transmitted Infections (IUSTI) World Congress, held in September 2022 in Zimbabwe.

### First round

The first round (supplementary information 1) aimed to survey the participants for ideas on how GIFT could be used in STI/BV and HIV management in LMICs. Participants were provided with background information on GIFT, including which attributes were considered to be fixed (being a lateral flow immune assay, in a POC test format, using a lateral vaginal wall swab sample type, and detecting cytokine biomarkers of genital inflammation), and which were in development and potentially modifiable (choice of biomarker combinations, turnaround time in 10-20 minutes, sensitivity between 77-86% and specificity between 68-71% [depending on a trade-off of biomarker combinations], and cost of each test between USD 0.50-4.00).

We used qualitative, open-ended questions to allow participants to provide as many ideas as possible, which could then be refined and focused into a core set of strategies in subsequent rounds. The questionnaire was divided into five sections. In the first section, participant demographic data were collected, including the participant’s professional role, organisation, and country. Section 2 asked about the implementation of GIFT for asymptomatic women, followed by open questions in section 3 on where the GIFT device could be implemented within WHO syndromic management guidelines for women presenting with vaginal discharge (Figure 1).^15^ Participants were also asked at what points GIFT could be implemented within the syndromic management guidelines, and what barriers and facilitators of implementation may be, with six different population groups/scenarios presented. Section 4 asked about GIFT’s use in general, for example, as a screening or as a diagnostic tool, and implications for the test’s attributes. The final section allowed participants to provide any further comment or feedback.

**Figure 1.**
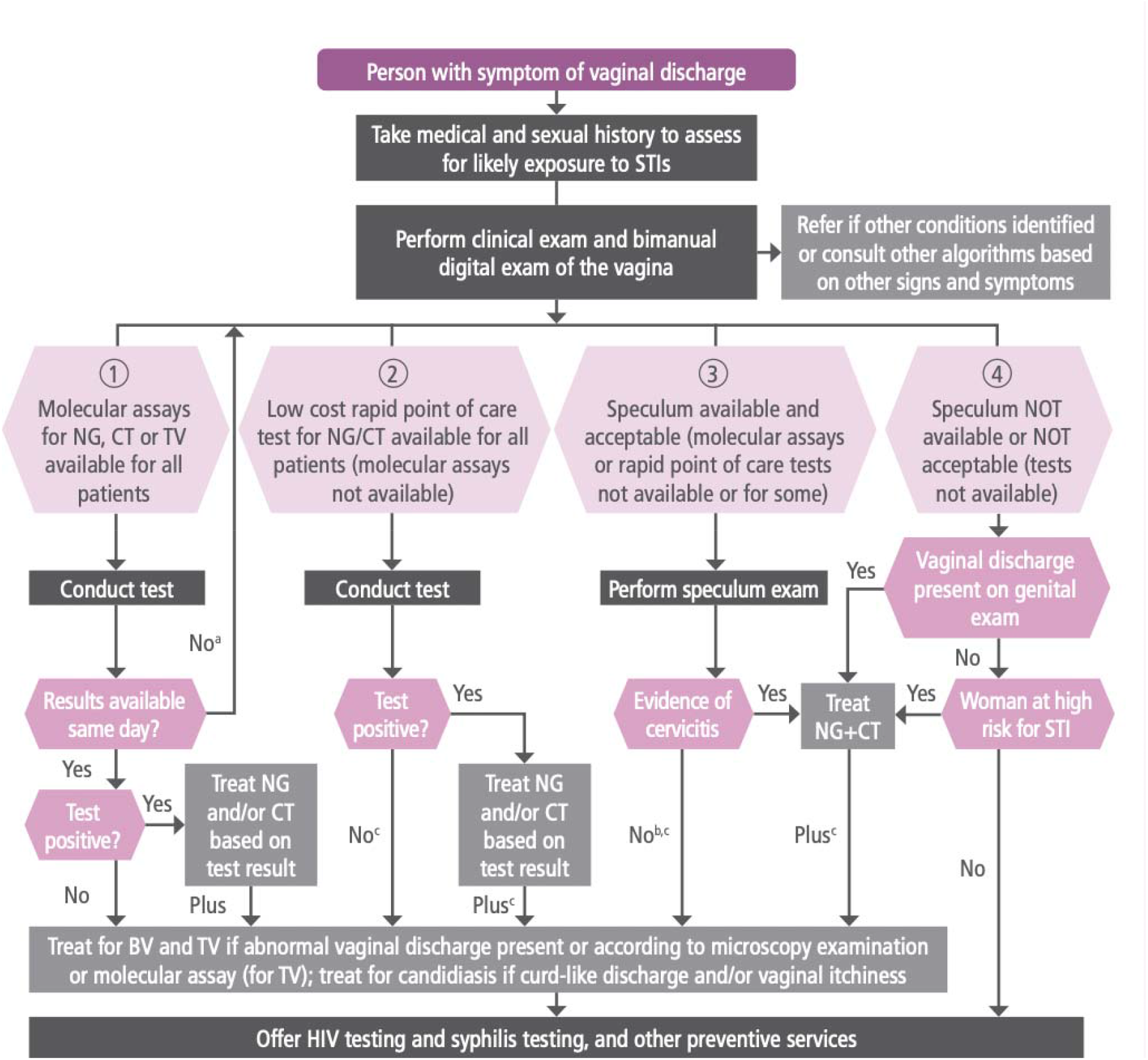
WHO guidelines for management of women with vaginal discharge^15^.

Quantitative data from close-ended questions were analysed using basic descriptive frequencies. Free text data from open-ended questions were analysed using thematic analysis, including coding text and identifying patterns and themes in the data.^19^

### Second round

The second round questionnaire (supplementary information 2) aimed to develop consensus on how GIFT could be integrated into STI/BV management algorithms, using the list of strategies emanating from the first round. Participants were asked to indicate their level of agreement for each strategy using Likert scales, ranking and multiple-choice questions, as well as opportunities for open-ended commentary. We asked participants to build consensus in the form of statements. We asked experts to consider the use of GIFT in symptomatic and asymptomatic women separately, as in the first round. As with round one, the quantitative data from close-ended questions were summarised in tables and the free text data from open-ended questions were analysed using thematic analysis including coding text and identifying patterns and themes in the data. For the purpose of this study, consensus was defined as general agreement of a substantial majority (≥70%) of participants.^20^ Consensus was reached on all statements in the second round questionnaire, and therefore no further questionnaire rounds were needed.

### Ethical considerations

Ethical approval was granted by the London School of Hygiene & Tropical Medicine Research Ethics Committee (Ref: 26626). An information letter and consent form were embedded in the survey forms (in both the online and offline forms) for participants to read and sign prior to completing the questionnaire.

## Results

A total of 28 responses were received in the first round and 68 in the second round (Table 1). Participants were largely researchers, clinicians and other healthcare professionals, with most practising in urban areas, rather than semi-urban or rural. In both rounds, there were more female than male participants, and the majority of participants came from the African region. In the first round, there were no participants in the 25-34 year age-bracket.

**Table 1:**
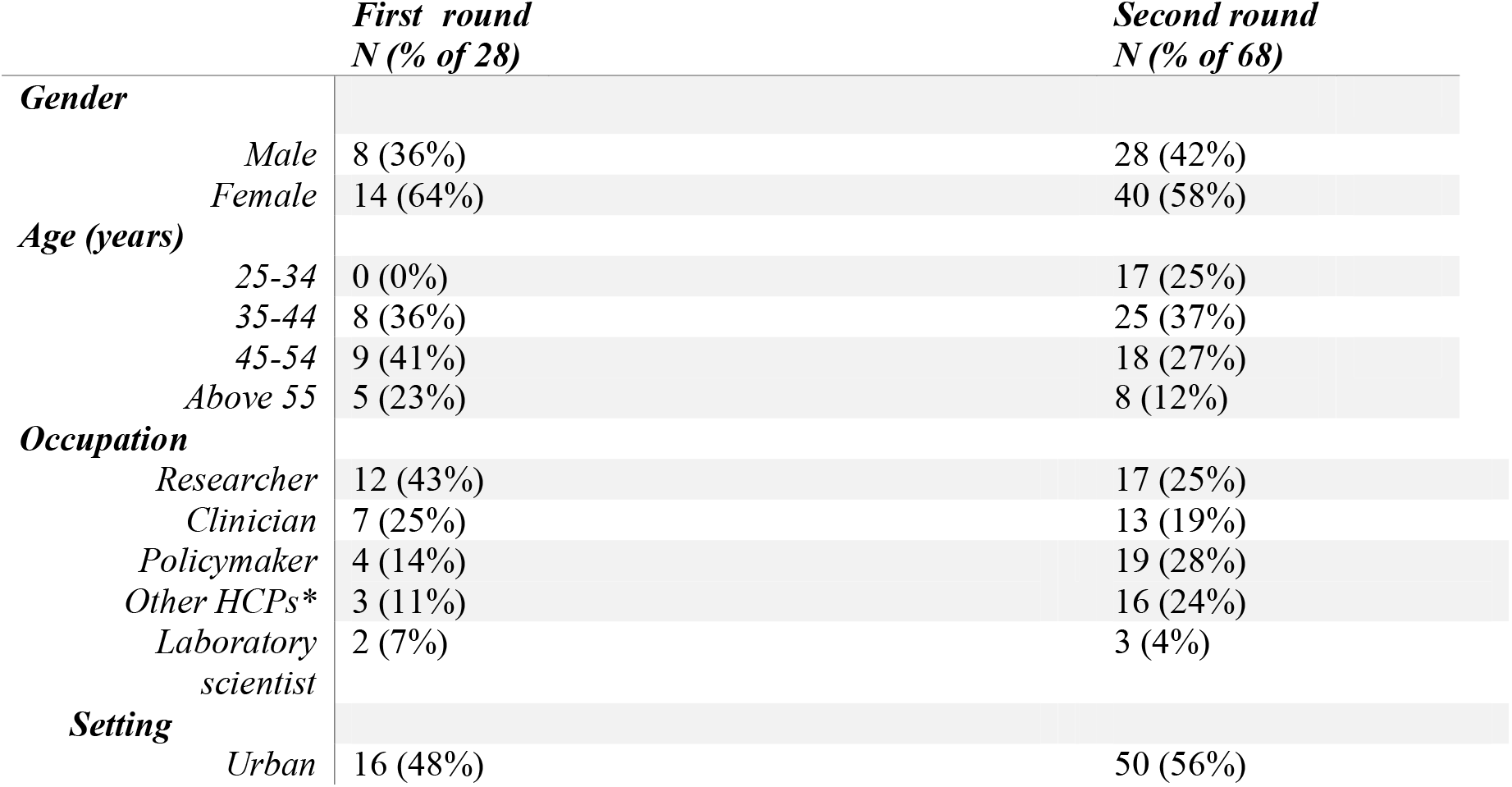
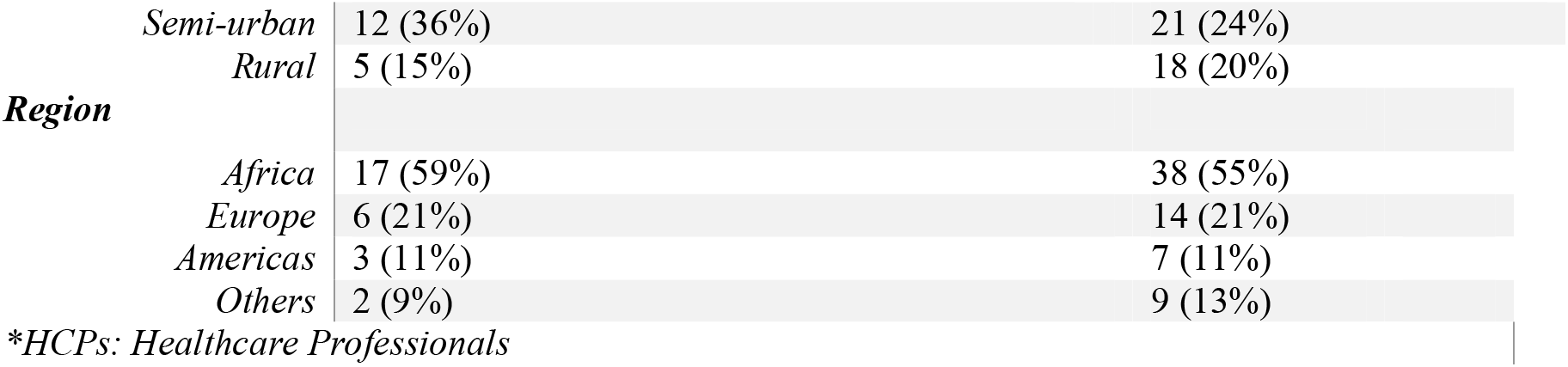
Participant socio-demographic characteristics for both rounds in the modified Delphi survey.

## Results from the first-round questionnaire

### Use of GIFT

Three-quarters (76%) of participants agreed that GIFT should be used as a screening tool i.e., a test that needs to be followed by aetiological testing. Participants explained that using GIFT as a screening tool was important, as many facilities in LMICs have little or no access to resources for aetiological testing, so this approach could triage access to testing. This could potentially facilitate case detection and linkage to care.

### Priority population groups among asymptomatic women

Eighty-nine percent of participants agreed that, most high-risk women (transactional sex or commercial sex workers) of any age should be highly prioritised for the use of the device out of six population groups proposed by the study authors (Table 2). The next highest-ranked group were young women and girls (16-24 years) and older (>=25 years) women in a high/moderate HIV prevalence setting. Additional groups suggested by the Delphi participants included women using pre-exposure prophylaxis (PrEP), those attending family planning clinics, and those attending cervical cancer screening clinics.

**Table 2:**
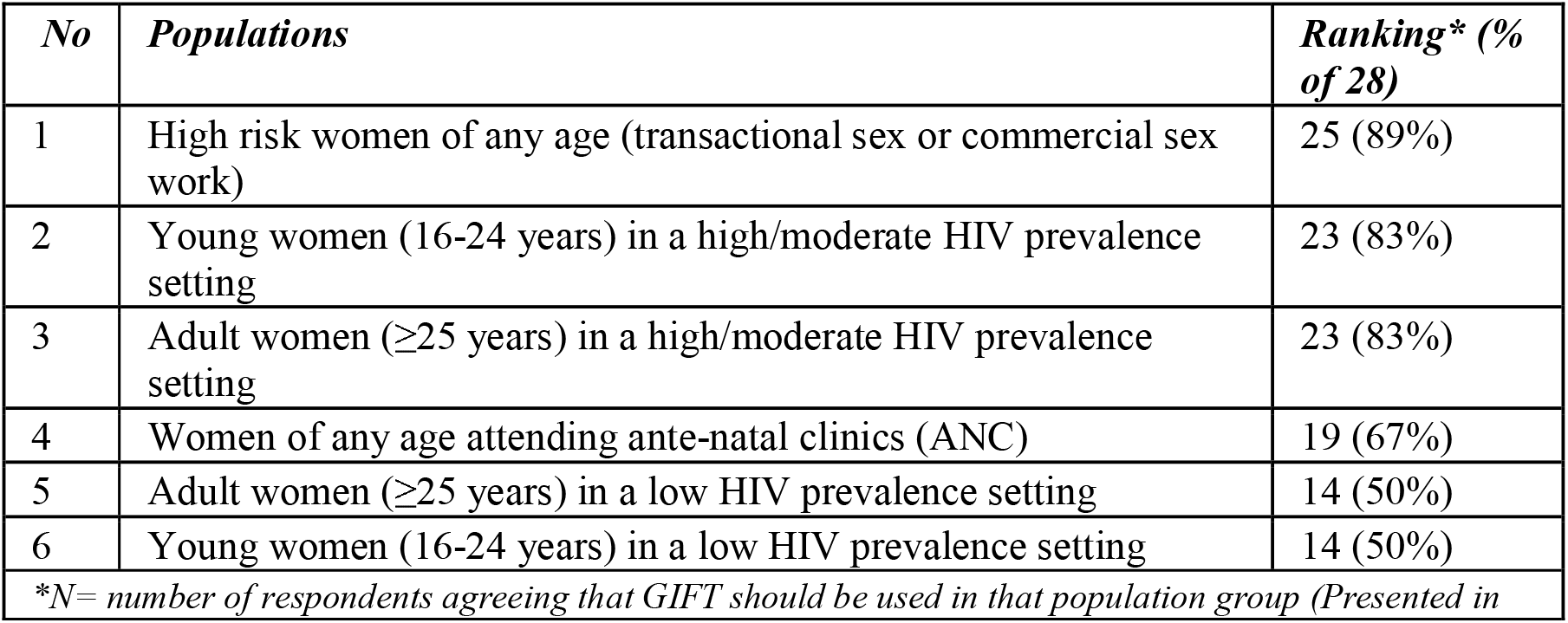
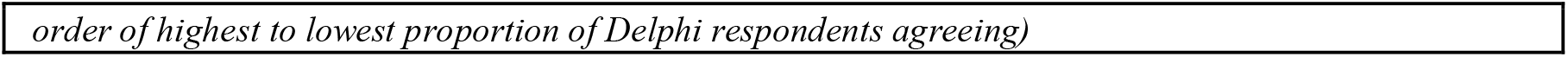
Ranking of asymptomatic population groups for GIFT use, based on the proportion of participants agreeing for GIFT use in the group.

| *No* | *Populations* | *Ranking\* (% of 28)* |
| --- | --- | --- |
| 1 | High risk women of any age (transactional sex or commercial sex work) | 25 (89%) |
| 2 | Young women (16-24 years) in a high/moderate HIV prevalence setting | 23 (83%) |
| 3 | Adult women ($\geq 25$ years) in a high/moderate HIV prevalence setting | 23 (83%) |
| 4 | Women of any age attending ante-natal clinics (ANC) | 19 (67%) |
| 5 | Adult women ($\geq 25$ years) in a low HIV prevalence setting | 14 (50%) |
| 6 | Young women (16-24 years) in a low HIV prevalence setting | 14 (50%) |

*\*N= number of respondents agreeing that GIFT should be used in that population group (Presented in*

### Integrating the GIFT device into current symptomatic STI/BV management guidelines

For symptomatic women with vaginal discharge, half of participants (51%) suggested GIFT could fit into pathways 3 and 4 of the WHO vaginal discharge management pathway (Figure 1)^15^, while 32% of participants picked either pathways 1 or 2. Pathway 3 describes a scenario where a speculum investigation is available and acceptable but molecular assays or rapid POC tests are not, whereas pathway 4 describes a scenario where a speculum investigation is not available or not acceptable, and other tests are also not available. The explanation for this was that GIFT would be best used as a screening test to be able to confirm inflammation during examination. Some of the participants (17%) reported the device needed a different pathway separate from those in the guidelines.

### Attributes and barriers associated with integration of GIFT into national guidelines

Common suggestions from free-text responses on how GIFT could be further improved to facilitate its integration into STI/BV management algorithms were: ensuring high accuracy (sensitivity and specificity), availability at all times, low cost and affordability, ease-of-use for end-users (whether they be healthcare providers or women themselves), ease of results interpretation, quick time to read results, and test stability. Cited barriers for integration included: lack of provider awareness, availability issues such as test stock-outs, high cost, and implications of test implementation (such as extra time and resources, including private space for sample collection that may be needed).

## Results of the second-round questionnaire

### Use of GIFT for asymptomatic populations

The majority of the participants (85%) indicated that they would offer GIFT to sexually active, asymptomatic women, with 69% indicating that a positive GIFT result should be followed by aetiologic diagnostic tests to confirm the cause of inflammation prior to treatment provision. When asked if GIFT could be used for a purpose other than screening, the majority (87%) said “No”. On a 5-point Likert scale from strongly agree to strongly disagree, ≥80% of participants agreed that GIFT could be used in each of the populations presented in Table 3.

**Table 3:**
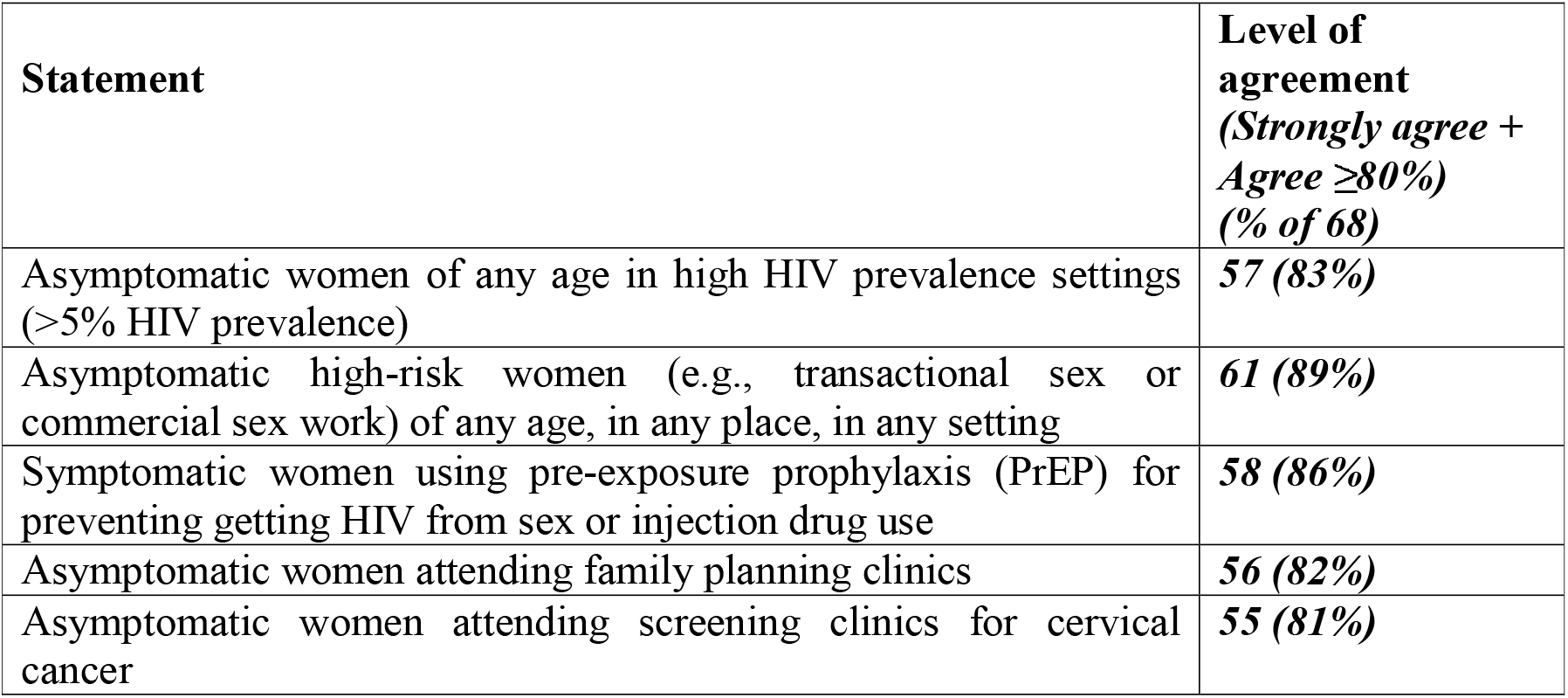
Asymptomatic populations for which Delphi participants agreed GIFT could be used.

### Integration of GIFT into current guidelines for STI/BV management among women with vaginal discharge

When asked at what stage would you introduce the GIFT device, respondents thought that the stages at which GIFT could be used within pathway 3 included: at the time of the vaginal clinical exam, alongside a speculum investigation to assess if there is evidence of cervicitis, and after observing evidence of cervicitis to assess whether to treat for an STI/BV. Regarding inclusion in pathway 4, 78% of respondents mentioned the time of vaginal clinical exam regardless of vaginal discharge presence, but only if the woman is at high risk for STIs/BV.

### Attributes and barriers for integrating GIFT into STI/BV management algorithms

All participants ranked all of the attributes that were identified in round 1 as either very important or important for integrating GIFT into STI/BV management algorithms. The attributes, ranked from most to least important (Table 5), included: affordability and costs, high sensitivity, specificity, PPV and NPV, fast turnaround time (results available in 10-15min), simple and easy to use, and storage (stable at room temperature).

**Table 5:** Proportion of second round Delphi participants considering attributes and barriers to GIFT integration into existing guidelines as very or moderately important.

| **Attributes listed by the participants** | ***Very Important or Moderately Important (>80%)***<br>***(% of 68)*** |
| --- | --- |
| Affordability and costs | **55 (81%)** |
| High sensitivity, specificity, PPV and NPV | **55 (81%)** |
| Fast turnaround time (results available in 10-15min) | **54 (79%)** |
| Simple and easy to use | **50 (74%)** |
| Storage (stable at room temperature) | **50 (74%)** |
| **Barriers listed by the participants** | ***Very Important or Moderately Important (>80%)***<br>***(% of 68)*** |
| Lack of test availability/stock-outs of test | **67 (99%)** |
| Implementation (added layer of complexity to existing algorithms) | **67 (99%)** |
| High cost | **66 (97%)** |
| Lack of provider awareness | **65 (95%)** |
| Private space to collect the vaginal sample | **62 (91%)** |

Barriers to integration of GIFT into existing guidelines for asymptomatic and symptomatic women identified in the first round were ranked as very or moderately important on the Likert scale in the second round, as presented in Table 5.

Additional considerations for GIFT implementation raised by the respondents in open text fields were: the need for awareness-raising and clear messaging among healthcare professionals and women, as well as appropriate training for healthcare professionals, given the novelty of the device and what it is measuring; and the potential for GIFT to be used as a self-testing device by women.

## Discussion

The Delphi survey results across the two questionnaire rounds with STI management programme and policy stakeholders suggest that the most likely use case for GIFT device is as a screening tool. The attributes prioritised for implementing the GIFT test in LMICs include ease of use, fast turnaround time, stability when stored at room temperature, affordability and high sensitivity and specificity. Participants also agreed that the device could be integrated into existing treatment guidelines using specified pathways. The main perceived barriers to its integration included test unavailability, complexities in its roll out, high cost, lack of provider awarenes and additional private spaces needed for the test within health facilities.

As expected, diagnostic accuracy was often considered a key attribute, ranked as highly important by most respondents. However, although diagnostic accuracy is important, it is not a sole criteria for successful test implementation, as there are many highly accurate tests already developed but never implemented into routine use.^12,13^ It is therefore important to consider the other attributes that would facilitate, or hinder, the tests implementation. Other highly-ranked attributes by respondents included low cost for the device to be affordable to national healthcare systems in LMICs, simple and easy to use, fast turnaround time to read results with easy interpretation, and not requiring any tailored storage conditions as it should be stable at room temperature. These attributes are consistent with the REASSURED criteria, which WHO considers to be the optimal attributes of POC tests.^21^ However, there is always a trade-off, as no test entirely meets all the criteria. It is therefore important to know which attributes are considered more important than others, in order to facilitate the implementation of a diagnostic; and this ranking of attributes may differ depending on context.^12^The prioritised GIFT attributes may be different if the test is to be used for a different use case, for example, as a diagnostic test rather than a screening tool. As part of the ongoing GIFT Africa study, a discrete choice experiment study is being conducted which will identify the trade-offs that participants (women attending the health facilities where the diagnostic study is being conducted) are willing to make between attributes when deciding whether to continue with STI management.^11^ These preferred attributes can be complemented by target product profiles (TPPs), used to help manufacturers prioritise test characteristics when developing tests, where depending on a test’s use case, the TPP characteristics may be different despite it being for the same disease.^22^ A draft TPP for GIFT has been developed by the study team, but there remains a need for formal development of TPPs for POC tests for genital inflammation and BV that follow internationally-recognised processes.^23^

We found that the main purpose of the GIFT device, given its current and predicted attributes, is its use as a screening tool to identify women who may have an STI and/or BV, with follow-up STI/BV testing to confirm aetiology prior to treatment. It was thought to have particular use as a screening tool among high-risk asymptomatic women, in-line with earlier studies that suggest rapid POC tests can improve the management and control of STIs in high-risk populations.^24^ The test would be particularly important for high-risk women who are asymptomatic, especially in LMICs where advanced diagnostics and testing resources are scarce. This finding is consistent with a recent study in Tanzania that found the prevalence of STIs among asymptomatic women in a birth control clinic to be high and called for routine screening tests and tools for these high-risk women in LMICs.^25^ However, a positive GIFT result would require a follow-up diagnostic test, supporting antimicrobial stewardship programmes to reduce empirical treatment and ensure antimicrobial drugs are used rationally. Given that the availability of additional tests is a recognised barrier to STI diagnosis in LMICs^7,15,26^ this may be a limitation to the acceptability of using GIFT as a screening tool if an aetiological diagnosis is not possible, but management based on a positive GIFT test is considered non-specific and prone to the same over-treatment challenges as syndromic management.^26^ This said, a third of participants felt that GIFT does not need to be followed by aetiological testing, as these are oftentimes not available in resource-constrained settings. In these scenarios, the presence of a POC screening tool such as GIFT would be valuable for management decisions. The GIFT Africa study also includes a qualitative study to assess the user experience, usability, and acceptability of the GIFT device at the POC, which will provide further, in-depth, insight into how GIFT could best be implemented to support patient management.^11^

In terms of integration into existing STI/BV symptomatic women management guidelines, consensus was reached that the device could be integrated into pathways 3 and 4 of the WHO 2021 guidelines for symptomatic women.^27^ This suggests feasibility and potential for utilising GIFT for management, especially in resource-limited settings where testing resources are lacking. The GIFT consortium is currently designing clinical algorithms integrating GIFT to optimise STI case finding and management.^11^ Earlier studies have reported that while many POC tests exist for STIs, there are implementation barriers at the level of the device itself, patients, providers, and the health system, hindering their use within treatment guidelines.^28^ It is important for test developers and healthcare managers to explore the feasibility of a POC test’s integration and its use case(s) during its developmental stages.^29,30^ Our study pre-defined certain use cases and population groups. The Delphi participants suggested additional use cases, such as for sexual health self-care, for HIV risk assessment to triage for PrEP referral, and for improving adherence to PrEP. The feasibility and acceptability of these additional use cases warrant further investigation, and the subsequent implications on priority device attributes and development should also be considered.

Our study has both strengths and limitations. We were able to recruit a diversity of experts with experience in STI/BV research and/or management in LMICs to provide feedback to inform the further development of the GIFT test and considerations for its implementation in routine STI management guidelines. We were also able to reach a consensus on all statements after two survey rounds, indicating general agreement on the test attributes and integration into management guidelines. Although our sample size was small, especially in round 1, we still reached data saturation for all topics covered in round 1, a more open-ended and qualitative, questionnaire. Another limitation is the fact that our study asked questions hypothetically, as the GIFT device is still in development, so it was difficult to develop concrete plans given that participants were not familiar with the test. However, surveying this stage means that results can feed into the final stages of device development.

The GIFT device has the potential to be a valuable POC screening tool for detecting genital inflammation. Further research utilising qualitative and mixed methods approaches on end user perspectives and preferences is needed to further explore successful integration into STI/BV management guidelines. Affordability of the device is key and stakeholder consultations will facilitate GIFT’s roll-out and sustained use within healthcare systems.

## Data Availability

All data produced in the present study are available upon reasonable request to the authors

## Author contributions

SF, LM and JAP conceived the study. SF, EHE and CMY developed the protocol. EEK, TC, KK, JvdW and EHE designed and tested the survey instruments. All co-authors disseminated the survey widely within their networks. EEK and EHE led the manuscript writing, with input from all other co-authors. All authors listed have read and approved the final version for submission.

## Acknowledgements

The authors would like to thank Professor Katharina Kranza for for reviewing and finalising the survey instrument and additional support with the dissemination to panellists. We also want to thank the Delphi participants for completing the survey and the members of the study Scientific Advisory Board (SAB) for providing feedback on the questionnaire design and for supporting the dissemination of the questionnaires within their networks.

## Supplementary files

Appendix I: Delphi survey first round questionnaire: https://form.jotform.com/220447010116035

Appendix II: Delphi survey second round questionnaire: https://eu.jotform.com/build/221927653855365

